# Association between preference and e-learning readiness among the Bangladeshi female nursing students in the COVID-19 pandemic: a cross-sectional study

**DOI:** 10.1101/2021.06.20.21259188

**Authors:** Humayun Kabir, Tajrin Tahrin Tonmon, Md. Kamrul Hasan, Lila Biswas, Md. Abul Hasnat Chowdhury, Muhammad Didarul Islam, Mamunur Rahman, Dipak Kumar Mitra

## Abstract

**Background:** The COVID-19 pandemic jeopardized the traditional academic learning calendars due to the closing of all educational institutions across the globe. To keep up with the flow of learning, most of the educational institutions shifted toward e-learning. However, the students’ e-learning preference for various subdomains of e-learning readiness did not identify, particularly among the female nursing students’ for a developing country like Bangladesh, where those domains pose serious challenges.

**Results:** A cross-sectional study was conducted among the female nursing students’ perceived e-learning readiness in subdomains of readiness; availability, technology use, self-confidence, and acceptance. The findings of the study revealed that the prevalence of preference for e-learning was 43.46%. The students did not prefer e-learning compared to ‘prefer group’ has significantly less availability of technology (β = -3.01, 95% CI: -4.46, -1.56), less use of technology (β = - 3.08, 95% CI: -5.11, -1.06), less self-confidence (β = -4.50, 95% CI: -7.02, -1.98), less acceptance (β = -5.96, 95% CI: -7.76, -4.16) and less training need (β = -1.86, 95% CI: -2.67, - 1.06). The age, degree, residence, parents’ highest education, having a single room, having any eye problems were significantly associated with the variation of availability of technology, use of technology, self-confidence, acceptance, and training need of e-learning.

**Conclusions:** The outcomes of the study could be helpful while developing an effective and productive e-learning infrastructure regarding the preparedness of nursing colleges for the continuation of academia in any adverse circumstances like the COVID-19 pandemic.

## 1. Background

E-learning is addressed as web applications that enable the participation of individuals either distinctively or synergistically, involving collaborative digital medium and virtual classrooms to reciprocate lessons via online settings (Padalino and Peres 2007). Such pedagogical structure accedes the mentee to stay hooked asynchronously with academic learning activities continue with the access of proper internet supplementation into the adapted devices (Horiuchi et al. 2009). Worldwide, healthcare educators were seen adapting this evolutionary method of education strategically with the help of continuously developed technologies as a distant instructional teaching component to share their knowledge and skills around the health communities (Beeckman et al. 2008). This required a fundamental shift of electronic device and internet-based teacher-learner reciprocity from the conventional physical presence-based education deliverance strategy (Horiuchi et al. 2009).

Both locally and internationally, increased interest in nursing educational programs continuation via e-learning is seen accelerating due to the concurrent need of timekeeping in mind the prime contents: quality, social distancing for emergency health occurrences, time flexibility, and cost-effectiveness (Sheen et al. 2008). In nursing, e-learning, when were accessible through hospital websites for the nurses, allowed them to widen their knowledge and skills by taking their required courses as the service deliverance heavily depends on their enriched cognizance. Besides, available nursing care information through the hospital websites implements the healthcare organizations to renovate professional and personal growth among the nursing community (Gega et al. 2007). The widely evident term e-learning facilitated the nursing learning system since the 1960s, according to the findings of the CAL (Computer-assisted learning) studies in the nursing literature where debates were persistent about the consolidated skill accretion of the nurses and the proficiency of conventional teaching methods within the clinical environment (Bloomfield et al. 2010). Ironically, e-learning had a greater drop-out rate than the traditional delivered education, despite the advantages, according to another study findings, due to the lack of computer competency, browser handling, unavailability of adequate technologies, and nursing students’ acceptancy towards it (Mohamed Ali 2016). E-learning concentrates on three parts enormously, including the appropriate words used for the presentations, particularly the networking medium used and the pedagogic motive to bring constructive changes in people’s lore (Mayer 2019). This learning system continued to grow with the growing interest of both faculty and pupils apart from the technical support upliftment with helping in courseware delivery (Beqiri et al. 2009). These online course delivery models can be both coeval and nonsynchronous (Chen 2016). Regardless of the models, e-learning was adhered to globally as the timely educational plan of action to adopt during the COVID□19 pandemic caused by SARS-CoV-2 in medical and nursing education due to the ongoing campus closing period (Lahti et al. 2014; Fawaz and Samaha 2020; Hossain et al. 2021).

Though dissensions are present regarding the e-learning educational system, by ensuring the accessibility, affordability, and flexibility of this systems’ teaching disquisition from both the disseminators’ and receivers’ sides, the learning capacity of the students can be developed for life-long study purposes (Dhawan 2020). The learning process should be utilized in accordance with the learners’ necessity and capacity to accommodate, assess and contemplate the instructors’ recitation (Heo and Han 2018). Students should be prioritized to be exiled to perform well while using the web-based educational method (Lahti et al. 2014). Although, it was found in a study that despite the training, e-learning could be depressive and stressful for visually impaired students (Lee and Oh 2017). To avoid these problems, specifications and standards using contents present in the LMS (Learning Management System) need to be created and accessed for the disabled people to access all the information given and overcome the sufferings (Guenaga et al. 2004). Similarly, a study observed that the quality of the training programs needs to be improved, and to develop the learning experience of the nursing students, training on generalized caregiving skills, knowledge, and self-efficacy of mental health should be provided (Irvine et al. 2007). From different points of view, in several studies, gender differences played some roles in perceiving e-learning acceptance (Ong and Lai 2006; Ramírez-Correa et al. 2015).

Moreover, perceived usage experience, the intention to learn, and the benefit of technology-based learning were found to be slightly lower among males than females (Ramírez-Correa et al. 2015). Similarly, a mixed-method study finding in Saudi Arabia, Mutambik et al. suggested gender divide investigations of’ e-learning readiness in deferent cultural setting (Mutambik et al. 2020). In addition, Bangladesh Nursing and Midwifery Council (BNMC) allows 90% of its seat for female students during admission.

In Bangladesh, very few studies were found investigating the barriers on its way to cope with the e-learning methods, which were not directly reporting the current situation of students’ readiness towards the e-learning system and their preference for this reason. The recent pandemic has taught us to be prepared for all the time, especially to be skilled and efficient enough to continue the educational and professional activities virtually in case of emergencies. However, there was no baseline research on female nursing students. For the continuation and building of a more sustainable nursing education system, in the current situation of a developing country like Bangladesh, e-learning readiness assessment is highly essential. Therefore, our study intended to explore the association between the preference and various domains (availability of technology, use of technology, acceptance, self-confidence, and training) of e-learning readiness among the Bangladeshi female nursing students with the hope to contribute to the educational system development to find newer tactics to deal with the found barriers in the steps of existing evidence. To achieve the aim of this study following research questions were articulately constructed:

1. What is the prevalence of e-learning preference among the female nursing students of Bangladesh?
2. Is there any association between preference and e-learning readiness subdomains among female nursing students?
3. What are the other variables associated with female nursing students’ e-learning readiness subdomains?

## 2. Methods

### 2.1. Study design

A cross-sectional study was conducted between December 26, 2020, and January 11, 2021.

### 2.2. Study participants

The study participants were all undergraduate female nursing students in Bangladesh. To reduce the recall bias, the inclusion criteria was the student who enrolled in e-learning at least in the last 30 days of this study period.

### 2.3. Data collection

Data were collected online using “Google Form,” posting the questionnaire link on nursing students’ social media groups (Facebook, Messenger, and WhatsApp) during the school closing period in Bangladesh due to the COVID-19 pandemic. Given the current situation, we followed convenience and snowball sampling methods and found 252 responses. Finally, a total 237 of completed responses were recruited for the analysis. A workflow describing the methodology of the research is presented in **Figure 2**.

**Figure 1.**
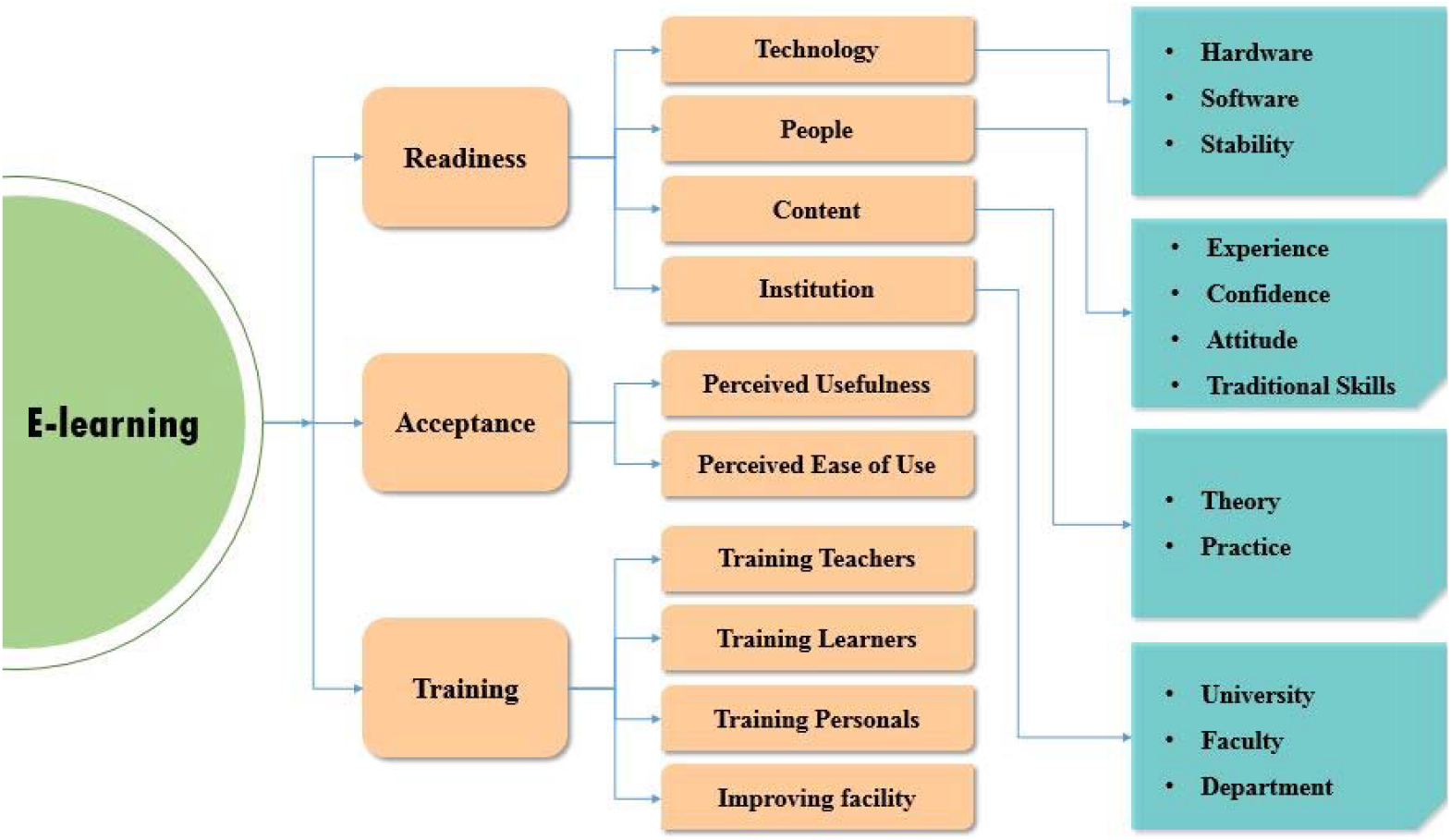
A Model for assessing Students’ Readiness for E-learning

**Figure 2:**
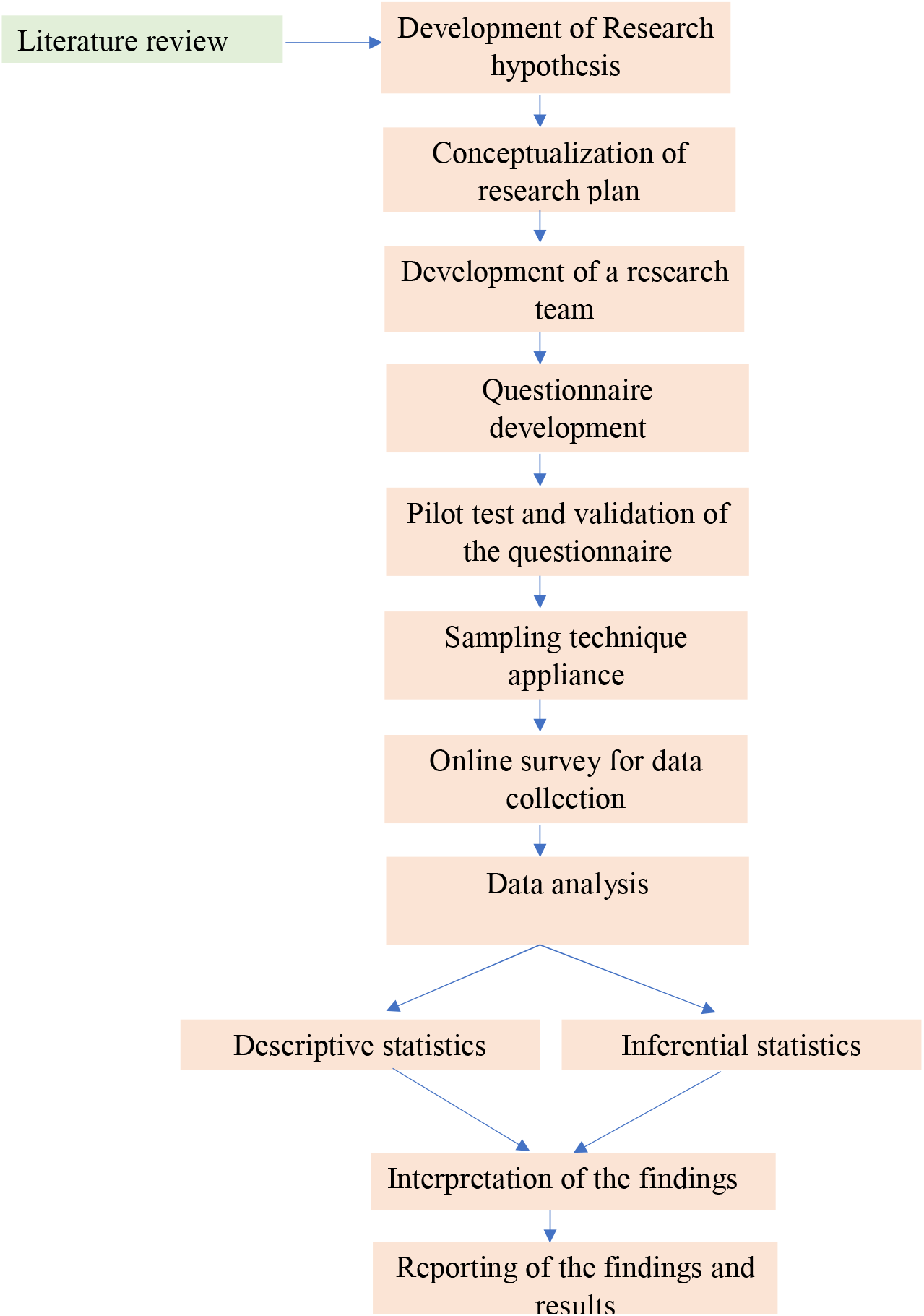
Workflow of the study.

### 2.4. Questionnaire development

The questionnaire included whether the students were willing to participate in the study, an item on perhaps the students prefer e-learning, other variables, and a perceived e-learning readiness questionnaire. The first part of the questionnaire included an item on students’ willingness to participate in the study. The second part consisted of preferred e-learning, device use, a single room, eye problems, and demographic information (age, type of institution, degree, residence, and parent’s highest education). The preference of e-learning was accessed by binary (yes and no) response to a single item. Having any eye problems has been defined in the study; the student could not stay on screen longer. However, the third part of the questionnaire consisted perceived e-learning readiness questionnaire.

### 2.5. Measurement of perceived e-learning readiness

The readiness of e-learning can be accessed by using 39 items of the perceived e-learning readiness questionnaire (score range: 39-195) (Ünal et al. 2014). A Model for assessing Students’ Readiness for E-learning is presented in **Figure 1**. Numerous studies assessed university students’ e-learning readiness using the tool previously (Akaslan and Law 2011a, b; Soydal et al. 2011). The questionnaire items were responded to a five-point Likert scale of 1 for “strongly disagree” and 5 for “strongly agree.” The questionnaire primarily focused on five baseline subdomains of e-learning; availability of technology for e-learning (6 items), use of technology for e-learning (11 items), the self-confidence of e-learning (12 items), acceptance of e-learning (7 items), and training of e-learning (3 items). In our study, we considered all of the five subdomains of the e-learning readiness questionnaire. The probable score range of the questionnaire was 39 to 195, whereas our study found 36 to 192. This 39 items questionnaire showed excellent reliability in our study (Cronbach alpha= 0.94). None of the reliability coefficients (Cronbach alpha) of the subdomains were found less than 0.76 in this study are presented in **Table 3**.

### 2.6. Data analysis

Descriptive statistics were performed for all variables. The perceived e-learning readiness questionnaire score was presented by mean, median, standard deviation (SD), and interquartile range (IQR). The reliability coefficient (Cronbach alpha) was calculated for the perceived e-learning readiness questionnaire. Multivariable linear regression models were fitted to find the association between students’ e-learning preference and e-learning readiness subdomains. We adjusted the models for participants’ e-learning preferences with other variables. The dependent variables were e-learning readiness subdomains; availability of technology, use of technology, self-confidence, acceptance, and training. The independent variable was the preference of e-learning, and all other variables were included as covariates. The p-value <0.05 was considered statistically significant at a 95% confident interval. Data were analyzed by using statistical software STATA-16.

### 2.7. Ethical issue

The Ethical Review Broad of the Faculty of Life Science, North South University, approved this study. The reference number is 2021/OR-NSU/IRB/0601. The aim and objective of the study were explained on the first page of the questionnaire, and the respondents who were willing to participate were considered as respondents of this study.

## 3. Results

### 3.1 Prevalence of preference and baseline characteristics of the study participants

The baseline characteristics of the study participants are presented in **Table 1**. In this cross-sectional study, 237 e-learning enrolled female nursing students were recruited. The prevalence of preference for e-learning was 43.46%. The median age of the participants was 21 (IQR: 20-22) years. Approximately 95% (n=226) of the participants were enrolled from private institutions and most of the students, 71.73% (n=170), were from a diploma in nursing background. The majority of the participants, about 59% (n=141), were from the rural areas of Bangladesh. About 54% (n=128) had higher secondary certificates (H.S.C.) Most of the participants, 63% (n=149), had no single room for e-learning. However, around 42% (n=101) of the respondents reported having any eye problems.

**Table 1:** Baseline characteristics of the study population (n=237)

| **Characteristic** | **n** | **% / IQR** |
| --- | --- | --- |
| **Prefer** |  |  |
| No | 134 | 56.54 |
| Yes | 103 | 43.46 |
| **Median Age in year** | 21 | 20 - 22 |
| **Age** |  |  |
| <20 | 29 | 12.24 |
| 20-22 | 156 | 78.06 |
| >22 | 52 | 21.94 |
| **Type of institution** |  |  |
| Private | 226 | 95.36 |
| Public | 11 | 4.64 |
| **Degree** |  |  |
| B.Sc. in Nursing | 67 | 28.27 |
| Diploma in Nursing | 170 | 71.73 |
| **Residence** |  |  |
| Rural | 141 | 59.49 |
| Urban | 96 | 40.51 |
| **Parents highest education** |  |  |
| Bachelor's & above | 41 | 17.30 |
| H.S.C | 128 | 54.01 |
| Primary | 68 | 28.69 |
| **Device use** |  |  |
| Personal computer | 6 | 2.53 |
| Handset | 231 | 97.47 |
| **Having a single room** |  |  |
| No | 149 | 62.87 |
| Yes | 88 | 37.13 |
| **Having any eye problems** |  |  |
| No | 136 | 57.38 |
| Yes | 101 | 42.62 |

Note: IQR = interquartile range

### 3.2. Descriptive statistics of the e-leaning readiness questionnaire

The descriptive statistics and Cronbach alpha of the subdomains of the e-leaning readiness questionnaire are presented in **Table 2**. The mean scores of availability of technology, use of technology, self-confidence, acceptance, and training were found 16.08 (SD: 5.90), 33.61 (SD: 7.75), 37.89 (SD: 9.94), 21.90 (SD: 22), and 10.76 (SD: 2.96), respectively.

**Table 2:** Descriptive statistics of e-learning readiness questionnaire (n=237)

| Scale | Mean | Median | SD | IQR | Cronbach alpha |
| --- | --- | --- | --- | --- | --- |
| **Availability of technology** | 16.08 | 15 | 5.90 | 12-21 | 0.84 |
| **Use of technology** | 33.61 | 33 | 7.75 | 29-38 | 0.77 |
| **Self-confidence** | 37.89 | 38 | 9.94 | 32-45 | 0.88 |
| **Acceptance** | 21.90 | 22 | 7.40 | 16-28 | 0.92 |
| **Training** | 10.76 | 11 | 2.96 | 9-13 | 0.76 |

### 3.3 Association between preference and female nursing students’ availability of technology

In **Table-3**, the results revealed that e-learning non-preferring students had significantly less availability of technology for e-learning (β = -3.01, 95% CI: -4.46, -1.56, *p* < 0.001) compared to preferring group. The older age group (> 22 years) was found significantly having more availability of technology (β = 2.73, 95% CI: 0.33, 5.14, *p* = 0.026) compared to younger age (< 20 years) students. However, compared to diploma degree, the students from B.Sc. degree were found to have less availability of technology significantly (β = -2.83, 95% CI: -4.47, -1.18, *p* = 0.001). The students enrolled e-learning from rural area compared to urban was found significantly having less availability of technology (β = -1.48, 95% CI: -2.86, -0.09, *p* = 0.037). The parents’ highest education bachelor’s and above compared to primary education was found to have more technology availability (β = 2.47, 95% CI: 0.29, 4.46, *p* = 0.027). On the other hand, not having a single room was shown to have significantly less technology availability (β = -2.01, 95% CI: -3.50, -0.53, *p* = 0.008).

**Table 3:**
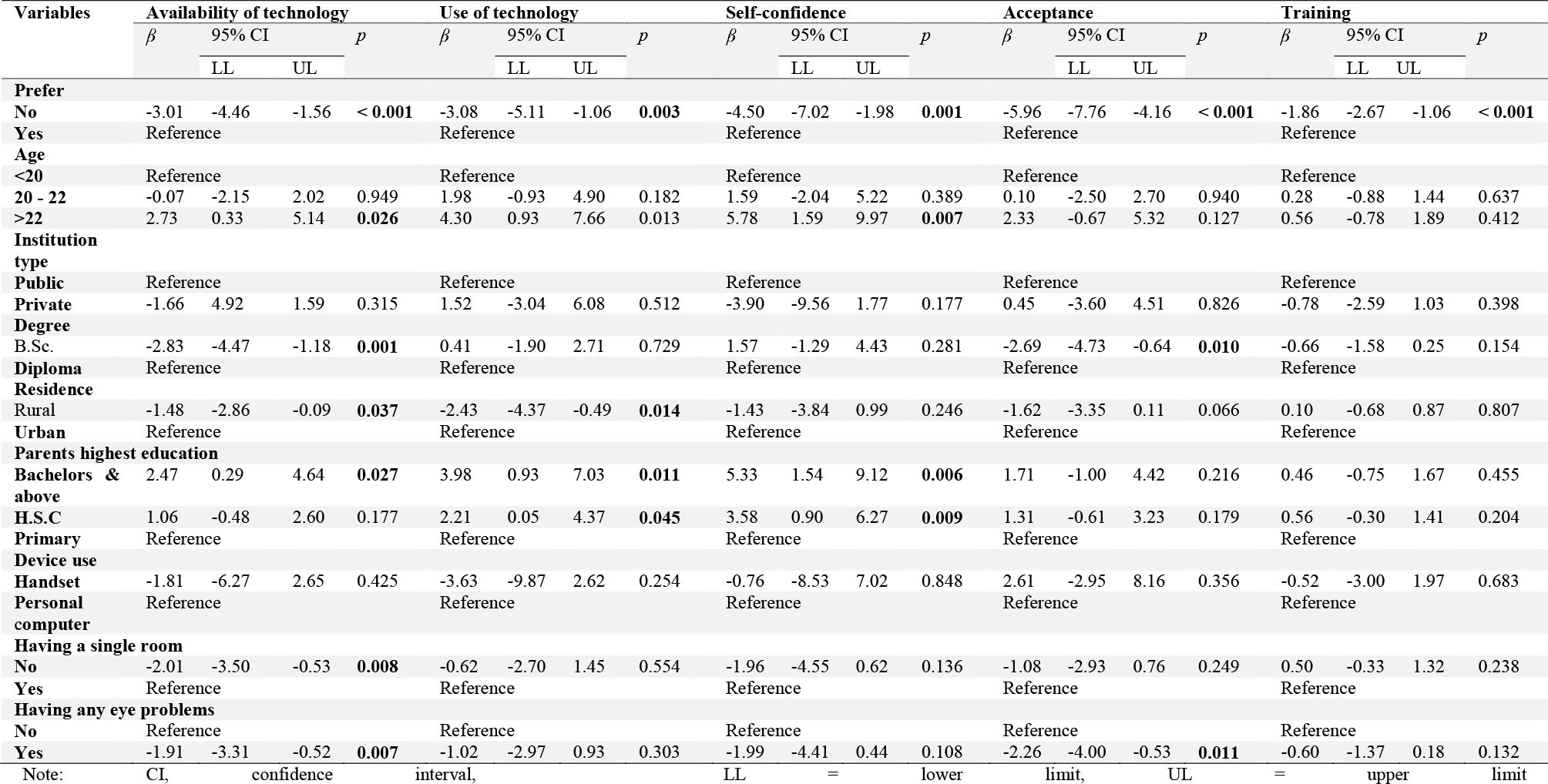
Association between e-learning preference and e-learning readiness and with other variables (n=237)

### 3.4 Association between preference and female nursing students’ use of technology

In this study, compared to prefer group the students who non-prefer e-learning were found significantly less use of technology for e-learning (β = -3.08, 95% CI: -5.11, -1.06, *p* = 0.003) (in **Table-3**). The older age group (> 22 years) was found significantly having more use of technology (β = 4.30, 95% CI: 0.93, 7.66, *p* = 0.013) compared to younger (< 20 years). The urban students were found significantly having less use of technology (β = 3.98, 95% CI: 0.93, 7.03, *p* = 0.014) compared to rural. The parents’ highest education bachelors and above was found significantly having more use of technology (β = 3.98, 95% CI: 0.93, 7.03, *p* = 0.011). Similarly, parents’ highest education H.S.C. compared to primary education found significantly having more use of technology (β = 2.21, 95% CI: 0.05, 4.37, *p* = 0.045).

### 3.5 Association between preference and female nursing students’ self-confidence

In **Table-3**, self-confidence of e-learning (β = -4.50, 95% CI: -7.02, -1.98, *p* = 0.001) was found significantly less among the non-preferring group compared to the preferring e-learning group. The older age students (> 22 years) were found significantly having more self-confidence (β = 5.78, 95% CI: 1.59, 9.97, *p* = 0.007). Significantly more self-confidence (β = 5.33, 95% CI: 1.54, 9.12, *p* = 0.006) was found among the students whose parents’ highest education was bachelors and above compared to primary education.

### 3.6 Association between preference and female nursing students’ acceptance

In **Table-3**, significantly less acceptance of e-learning (β = -5.96, 95% CI: -7.76, -4.16, *p* < 0.001) was observed among the non-preferring students compared to preferring e-learning students. The B.Sc. degree holders acceptance was found significantly less (β = -2.69, 95% CI: - 4.73, -0.64, *p* = 0.010) compared to diploma degree holders. However, less acceptance was found among having any eye problems students compared to not having (β = -2.26, 95% CI: - 4.00, -0.53, *p* = 0.011) any eye problems.

### 3.7 Association between preference and female nursing students’ training need

In **Table-3**, the results show that the non-preferring students’ training need was found significantly less (β = -1.86, 95% CI: -2.67, -1.06, *p* < 0.001) compared to the students who prefer e-learning.

## 4. Discussion

This study investigated the prevalence of e-learning preference and its association with the e-learning readiness domains and addressed the associated variables among the female nursing students of Bangladesh.

The study results showed that among all the participants, more than half of the students did not prefer e-learning. When we tried to figure out the factors that may affect this prevalence, it was observed that less acceptance and lack of self-confidence are two significant reasons for non-preferring e-learning among female nursing students. In addition, lack of technology usage, non-availability, and lack of training was also significantly associated with the non-preference. In this study, age differences, degree variation, residency, not having a single room while having online classes, and having any eye problems are also evidently associated with the non-preference among the female nursing students. In this study, 97.47% of the students used a handset (mobile phone, tablet) for e-learning. Albeit, availability of advanced technology is an integral part of e-learning, a study found, in a developed country (Hong Kong), during the COVID-19 pandemic, slightly more than half (66.6%) of the students attending online class via desktop or laptop and only 22.4% via mobile phone (Ho et al. 2021). Therefore, it can be said that the availability of technology is widely varied in developing versus developed countries.

The results of this cross-sectional study are consistent with earlier reports showing the effects of a higher frequency of not adopting e-learning due to several reasons. Those are readiness and other technological factors that significantly affect the study population. The perceived readiness of mind contemplates a person’s capability to inculcate e-learning in themselves (Al-Amin et al. 2021). Thus, a learner’s acceptance of using the newer technology-based method and the person’s self-efficacy is the impactful factors affecting the growth of e-learning. In this study, the students’ perception of readiness was reported similarly following other studies conducted in different circumstances, confirming this significant association claim (Mohamed Ali 2016; Bigirwa et al. 2020).

A study conducted in the context of the effectiveness of e-learning in Bangladesh showed that e-learning is a valuable system for the students’ pedagogical development. Nevertheless, the students’ perception of the lack of acceptance, technological efficacy, and motivation could hinder this development. It is one of the prime indicators of our study investigating the factors affecting the female nursing students’ readiness to adopt e-learning (Ali et al. 2018). A systematic review on nursing education revealed that students with higher satisfaction and performance were more prone to acceptance of web□based education (Du et al. 2013). Similarly, studies found that self-efficacy was significantly positively correlated with the success of e-learning (Yukselturk and Bulut 2007). In this study, the non-preference group showed a lower level of self-confidence toward e-learning.

Along with a high-performance score, a higher level of self-efficacy was observed among nursing students (Rouleau et al. 2019). Alongside, motivational training intervention fine-tuned their positive attitude toward e-learning (Rouleau et al. 2019). Henceforth, the current study found lower training needs among the female nursing students with non-preference of e-learning. However, the research found that training skills and ICT (Information Communication and Technology) metacognition skills improve learners’ level of achievement (Zimmerman et al. 1994; Salehi et al. 2014; Abdelrahman 2020).

To digitalize Bangladesh, the internet-based education system is expanding country-wide, but overseeing the access to the information and the programs, lagging behind the female nursing students of the nation to be more familiar with the system, which could be causing the lackings in preparing themselves to prefer the e-learning system more enthusiastically. A study conducted in Vienna also reported similar problems with e-learning adaptation similar to this study’s findings (Coopasami et al. 2017). This study showed that students aged more than 22 years were more inclined to prefer e-learning as they were more confident and had the technological support more available than the younger students. Wherewith slight inconsistency, H. Pillay et al. reported that students more than 40 years were with less self-efficacy and technological skills, and the younger age groups less than 25 years possess higher capability on both of these constructs (Pillay et al. 2007). From our country’s perspective, it could be explained that as more the students become experienced and exposed to this newer technology, they tend to use more of it. Hence this group will find it more available.

In this study, the association was found significant between educational degree variation and the lack of readiness in all the subdomains except for technology and self-confidence. It showed they (B.Sc. degree) have a lack of availability of technology and acceptance instead. Several studies around the globe on e-learning preference and readiness also found a significant association with the concerned factors (Smith 2005; Wei and Chou 2020). Similarly, a study conducted in the same context among the midwives learners had shown significant differences where the higher degree holders with more experiences showed more tendency to accept the e-learning than the junior degree holders (Ngampornchai and Adams 2016). In addition, during the COVID-19 pandemic, routine academic yearly examinations of the diploma degree were conducted by BNMC like the one the previous years based on e-learning.

On the contrary, the B.Sc. degree is run by public universities, and based on e-learning assessment, no yearly examinations have been conducted yet ([CSL STYLE ERROR: reference with no printed form.]). Therefore, lower e-learning acceptance might be found high among the B.Sc. degree students, which might explain why the academic year muddled up. However, the lower e-learning acceptance might be declined their perceived availability of technology.

A relatively higher tendency of the students to use smartphones was found to be more prone to accept online learning in the USA and India studies. The present study differs from those studies and could not find any significant association between preference and e-learning readiness according to availability, use of technology, and self-efficacy. In addition, the factor ‘device use’ has no association with readiness (Kobayashi 2017; T et al. 2020).

Rasha A. examined that environment affects learning (Raman 2016). Our study has not found any significant association between having a single room and its effect on the lack of e-learning readiness. However, it showed that those who were not having a single room lacked technology availability. Similary, Ivwighreghweta et al., (2014) observed that most participants in Nigeria prefer to access e-learning in a quiet and calm environment like cybercafes (Ivwighreghweta and Igere 2014).

This study also suggested that the lack of readiness for e-learning is also, to some extent, significantly associated with the residency of the female nursing students. In a similar study conducted in India, T. Muthuprasad et al., (2020) reported that the majority of the students living in a rural setting are associated significantly with the lack of readiness due to unavailability and self-efficacy, and another study also found the residency to be a potential factor in this regard (Elnakeeb et al. 2016). The advancement of technology and availability of it, with the speed it has reached the urban areas, did not find reaching in rural settings. Thus, the students studying from a rural setting perceiving online education are not privileged with available technical support and thus not enough confidence to use it. It was shown in the present study that the types of the university did not have any significant association with readiness. At the same time, a study in Kenya had also shown similar outcomes by reporting that both private and public universities were adopting the e-learning technique for educational deliverance (Neema-Abooki and Kitawi 2014).

However, in this study, having any eye problems showed significant association with having lack of availability of technology and being less acceptant towards the e-learning preparedness. A study addressing the problem faced by the disadvantaged people similarly showed that e-learning readiness, including self-confidence, acceptance, and technical availability, influenced their preference for technology-based learning (Hsieh et al. 2008). Insufficient instructional study for disadvantaged people, including people having physical discomfort like having eye problems in general, makes it difficult for them to endorse the technology-based pedagogy.

A significant number of participants in this study did not prefer e-learning, and their readiness was low. Thus, after completing the degree, ‘up to the mark’ professional development will be questioned. Our study finding is supported by Ho et al., who found that students’ competence in technology predicted e-learning preference significantly during the COVID-19 pandemic (Ho et al. 2021). Therefore, strategies should be implemented to strengthen educational policies regarding the e-learning readiness of the students. Similarly, e-learning readiness should be examined broadly by other developing countries worldwide in considering students’ future professional development and to mitigate potential learning gaps due to the ongoing pandemic.

## 5. Strength and limitation

Very few studies were found conducting the effectiveness of the e-learning for the Bangladeshi students, where no such studies were explicitly found investigating the efficacy of this method among the female nursing students of Bangladesh so far, which is a strength of this study. However, a study was found reporting the effectiveness of online learning, and the study considered a limited number of variables where the factors affecting the readiness for e-learning systems were not covered with a wide range of variables.

Although a readiness evaluation is essential, this research only highlighted the five aspects of readiness: the availability of technology, use of technology, self-confidence, acceptance, and training. As it was a cross-sectional study, the study could not investigate the range of its variables over a large group of female nursing students. Hence, it is recommended for future research to assess the various other readiness factors (sociological, environmental, human resource, financial, and content) on a larger scale study to report how ready the female nursing students of Bangladesh are to implement e-learning.

## 6. Conclusions

To reduce the spread of COVID-19, the closing of academic institutions and the introduction of e-learning were appreciated globally. Nevertheless, it may not be easy to assume that everyone is welcoming e-learning initiatives in a developing country like Bangladesh and ready enough. This study documented female nursing students’ preference for e-learning and its’ significant association between different subdomains of readiness in terms of availability of technology, use of technology, self-confidence, and acceptance. The study’s outcome showed that students’ preference has a significant association with the readiness of e-learning. The findings also suggested that other associated variables varied the e-learning readiness domains. This study’s findings might fill the gap of no baseline information about Bangladeshi nursing students’ e-learning readiness, particularly females.

## Data Availability

Data will be available on request to the corresponding author.

## List of abbreviations

COVID-19: Corona Virus-19

## Ethics approval and consent to participate

The Institutional Review Board (IRB) of North South University, Bangladesh, approved the current study. The purpose of the study was explained on the first page of the survey, and the respondents were asked on the first question whether they were willing to participate in the study and those selected ‘yes’ as written consent were considered to participate in the study.

## Consent for publication

Not applicable.

## Availability of Data Materials

Dataset used in this study will be available as per request (mailing to the corresponding author).

## Competing interest

The authors report no competing interest. The authors alone are responsible for the content and writing of this article.

## Funding

No funding from any public, private or non-profit research agency was received for this study.

## Author Contributions

Conceptualization, M.K.H., H.K.; methodology, H.K. M.K.H.; validation and scrutinization, D.K.M, H.K. M.K.H.; investigation, M.K.H., H.K.; writing-original draft preparation, H.K., T.T.T., M.K.H., L.B., M.A.H.C., M.D.I., M.R.; review and editing, D.K.M., M.K.H., H.K.; supervision, D.K.M.; All authors have read and agreed to the current version of the manuscript.

## Acknowledgments

We would like to accolade all the research assistants of the project who assisted in data collection.

## Notes

### Competing Interest Statement

The authors have declared no competing interest.

### Author Declarations

The Ethical Review Broad of the Faculty of Life Science, North South University, Bangladesh approved this study. The reference number is 2021/OR-NSU/IRB/0601. The aim and objective of the study were explained on the first page of the questionnaire and the respondents who were willing to participate were only considered as respondents of this study.

